# Sustained expression of inflammatory monocytes and activated T cells in COVID-19 patients and recovered convalescent plasma donors

**DOI:** 10.1101/2020.11.17.20233668

**Authors:** Ravinder Singh, Meenu Bajpai, Pushpa Yadav, Ashish Kumar Maheshwari, Suresh Kumar, Sonal Agrawal, Jayesh Kumar, Islam Md, Jaswinder Singh Mars, Gayatri Ramakrishna, Shiv K Sarin, Nirupma Trehanpati

## Abstract

Intense monocyte activation and infiltration into the target tissues is the main mechanism of lung injury in SARS CoV2 infection. A reduction in the degree and nature of such cellular responses is expected following recovery. We aimed to investigate the immune responses in severe Covid-19 patients and recovered patients.

**Methods:** Severe COVID-19 patients (n=34) at Lok Nayak Hospital, New Delhi and COVID-19 recovered patients (n=15) from mild disease and considered for convalescent plasma (COPLA) donation at Institute of Liver and Biliary Sciences (ILBS), New Delhi were recruited. We performed a multiplex cytokine bead assay in plasma and detailed multicolour flow cytometric analysis in peripheral blood of both groups and outcomes were compared in both groups and with healthy controls (n=10).

**Results:** A significant increase in inflammatory markers [MIP1-a, MIP3a, MCP1, MIF, MMP12, ITAC, VEGF-A, and leptin] was observed in severe patients. Non-survivors additionally showed increased IL-6 levels. Despite the sustained expression of MIPs, the recovered patients showed a surge in MCSF and IL-18 levels. Both the groups had increased CCR2, CX3CR1 positive monocytes, low CD8 T cells, APRIL and BAFFR^+ve^ B cells compared with healthy subjects. In conclusion, patients who have recovered and considered for COPLA donations still have compromised immunity with sustained expression of inflammatory monocytes and activated T cells.

## Introduction

In December 2019, a new member of Coronavirus family emerged in Wuhan, China; later termed as Severe Acute Respiratory Syndrome (SARS) associated CoV-2 (Zhou et.al 2020, Chen et al 2020, Jiang et.al2020) causing a severe form of respiratory illness which spread across the globe involving more than 12 million people with almost 0.6 million deaths In India, SARS-CoV-2 has infected around 6.69 million patients and has claimed more than 1.04 million lives. The clinical manifestations of COVID-19 varied from mild disease with fever, cough, and little or no pneumonia to severe disease which presented as dyspnoea, hypoxia, and pneumonia and finally to critical disease characterized by systemic shock, respiratory and multi-organ failure (Huang et al 2020).

SARS-CoV-2 is transmitted through the respiratory route; the initial site of infection for the virus is the respiratory epithelium. After infecting the lung epithelium, the inflammation spills over into the circulation generating an immune response mediated by leukocytes and cellular mediators of innate immunity including monocytes, macrophages, and dendritic cells (DCs). Indeed, SARS CoV2 may infect the circulating monocytes and blood-derived macrophages (Zhang et al 2020). Infected monocytes differentiate into macrophages with a greater expression of chemokine receptors and traffic via chemo attractants to the target organ. Stimulation of these receptors induces the expression of pro-inflammatory cytokines. Most of the COVID-19 patients exhibit lymphopenia and pneumonia with a higher levels of cytokines in severe disease indicating that the virus-induced exacerbated immune responses play a key role in the pathogenesis, morbidity, mortality and recovery in patients. Further, T cells activation and excessive elimination of regulatory T cells add to uncontrolled pathogenesis in COVID-19(Qin et al 2020, Fung et al 2020).

There is no targeted drug therapy for COVID-19 available at present, although some studies have indicated benefits with, intravenous remdesivir, a combination of lopinavir and ritonavir, and also with convalescent plasma therapy (Cao et al 2020, Grein et al 2020, and Shen et al 2020). Convalescent plasma therapy is considered very beneficial initially and still being considered by many groups (Chen et al 2020, Zhang et al 2020, and Kai Duan et al 2020). However, still there is a key question about durability and longevity of immunity in recovered patients. Therefore, **our aim** in this study was to analyse immune markers in COVID-19 patients with severe disease and in recovered patients (who had recovered from the mild form of the disease and considered for convalescent plasma donations).

Here we described novel observation that increased activated monocytes, T cells, and low APRIL and BAFFR expressing B cells are present in COVID-19 patients and also have sustained existence of these markers in COPLA donors even after 3 weeks of recovery.

## Results

The mean age, gender, and body mass index (BMI) were comparable between the groups. The COVID RT-PCR+ve patients had severe disease with a high respiratory rate (34.47 ± 2.5), low O2 saturation (85 ± 4.03), low baseline PaO2/FiO2 ratio (162.92 ± 13.77), and X-ray changes (Table.1). Recovered patients (with history of mild infection) who were RT-PCR negative and in whom at least 14 days had passed after recovery were considered for convalescent plasma donation (COPLA donors) and they formed the second group.

**Table: 1.** Baseline characteristics of COVID-19+ve patients, recovered patients as COPLA Donors and healthy subjects

| Base line parameters | COVID-19 Patients (n=34) | Recovered patients- (COPLA Donors) (n=15) | Healthy Subjects (n=10) |
| --- | --- | --- | --- |
| Mean Age (Y ± SD) | 48.21 ± 9.79 | 38.07 ± 7.7 | 48.14 ± 9.05 |
| Male (n, %) | 22 (75.86%) | 15(100%) | 7 (74%) |
| Chest X-ray changes (n, %) | 25 (86.20) | - | - |
| BMI ( Mean and SD) | 26.31 ± 2.29 | 26.84± 2.63 | 26.28 ± 2.52 |
| Respiratory rate/min ( Mean and SD) | 34.5 ± 2.55 | 14.14± 2.53 | 14 ± 2.65 |
| PaO2 (mmHg) | 61.76 ± 4.96 | - | - |
| O2 Saturation (%) | 85.03 ± 4.03 | 99±1 | 99 ± 1 |
| FiO2 (mmHg) | 0.38 ± 0.04 | - | -- |
| PaO2/FiO2 ratio | 162.92 ± 13.77 | - | -- |
| Baseline Neutrophils (N, median and range) | 3375 (2626, 6928) | 4328 (2789,6100) | 4535 (3160, 6000) |
| Baseline Lymphocytes (L median and range) | 968 (758, 1874) | 2371 (1400-2600) | 2453 (1550, 2890) |
| N / L ratio | 3.8(3.37, 4.67) | 1.81(1.6, 2.0) | 1.67 (1.52, 1.84) |
| Platelet count (in lakh) (cmm3) | 1.8 ± 1.20 | 1.6 ± 1.43 | 2.2 ± 2.42 |
| SOFA Score | 7.44 ± 1.86 | - | -- |
| Baseline Ct value of RT-PCR | 31.91 ± 3.44 | -ve | -- |

### Plasma Profile of COVID-19 patients and recovered patients (COPLA donors)

Principal component analysis revealed that three groups have differential expression of analytes (Fig. 1A). Heatmap showed that MIP1-a, MIP3a, MCP1, MIF, MMP12, ITAC, VEGF-A, and Leptin were significantly increased but Fractelkine, MIP1b, ENA78, IL-18,MCSF, E-selectin, IP1-0, Il-2, IIFN-g, IL-1B, Il-6, and Il-10 were decreased in COVID-19 positive patients compared to healthy subjects (FIG.1 A-B). Patients who recovered and donated their convalescent plasma(COPLA donors) after 15-21 days of recovery, still showed higher levels of VEGF-A, Leptin, MMP12, and MIP1a, but, there was a significant surge of MCSF and IL-18 which was not observed in COVID-19 patients.

**Figure 1.**
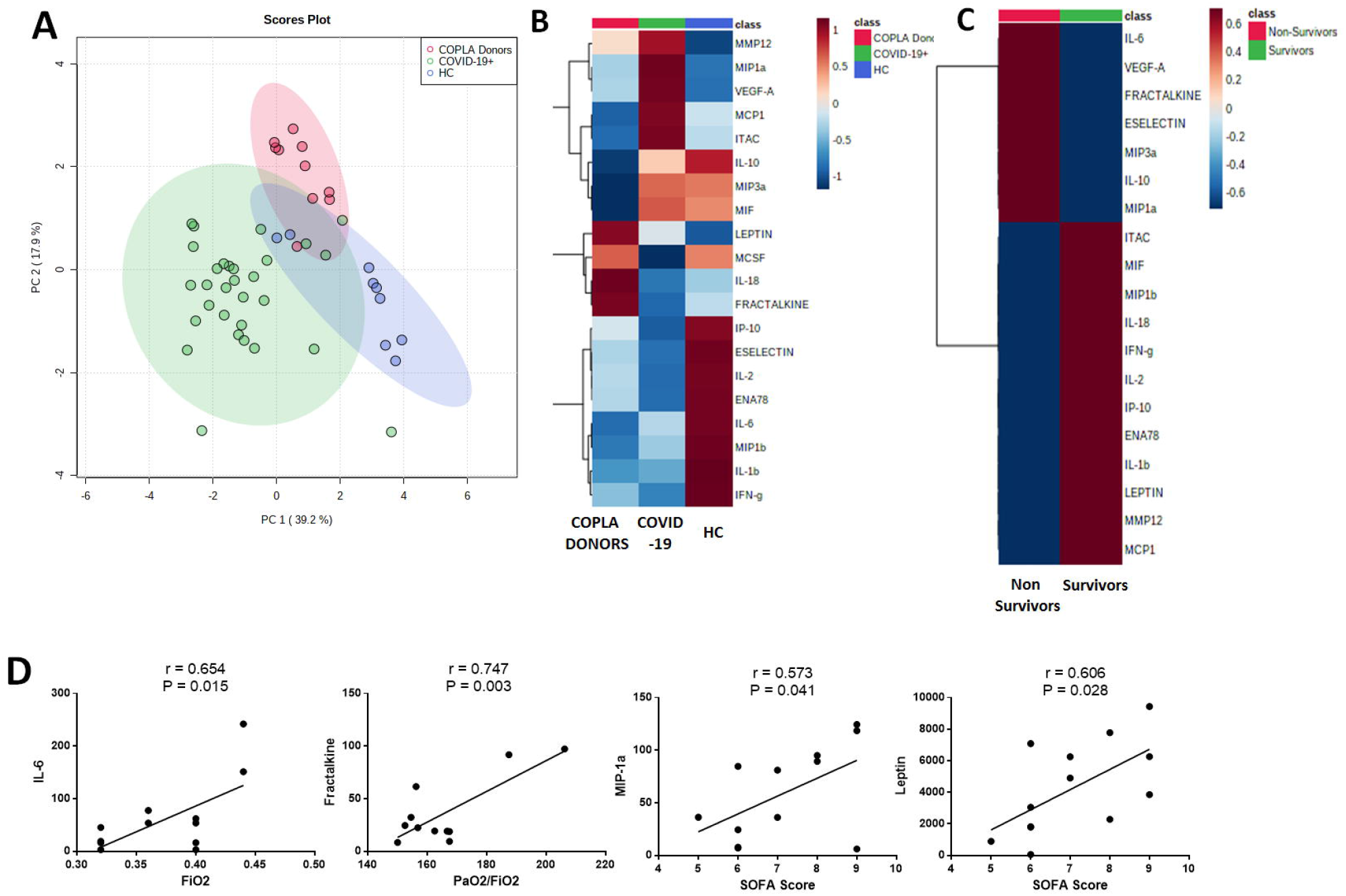
Differential expression of plasma analytes in COVID-19 patients, COPLA donors and healthy subjects. **(A)** Principal component analysis separating three groups. **(B)** Increased expression of MIP1-α, MIP-3α, MCP1, MIF, MMP12, ITAC, VEGF-A and Leptin in COVID 19 patients differentiate them from COPLA donors. COPLA donors had higher levels of VEGF-A, Leptin, MMP12 and MIP1a with additional surge of MCSF and IL-18.**(C)** Severe COVID-19 patients who did not survived had increased levels of IL-6, IL-10, MIP3a, Fractelkine and VEGF-A. **(D)** IL-6, MIP1-α, leptin and Fractelkine expression significantly correlating with Fio2 levels and SOFA scores. P value less than 0.05 was considered as significant.

COVID-19 patients who did not survive for a week due to severe COVID19, showed significantly increased IL-6, IL-10, MIP3a, MIP1-a, Fractelkine, and VEGF-A (Fig. 1C).

Finally, correlation analysis showed that few plasma analytes were significantly correlated with clinical parameters in COVID-19 patients. Increase in IL-6, MIP1β, and Leptin but decreased concentrations of Fractelkine was significantly correlated with FiO2 concentration and SOFA score (Fig. 1D).

### Increased CXCR3 and CCR2^+ve^ Monocytes in COVID-19 positive Patients and recovered COPLA donors

The total circulating monocyte population was increased in severe patients and recovered COPLA donors compared to healthy controls. Gating strategies of monocytes is given in supplementary material. Further analysis of monocyte distribution, based on CD14+CD16+ expression, we observed there was no significant change in the number of classical monocytes (CD14++16-ve) but an increase in intermediate (CD14++16+) and non-classical (CD14+CD16++) subsets in patients compared to healthy controls (FIG. 2). Despite no change in numbers of classical monocytes, this subset showed dual expression of HLA-DR/ CCR2 and HLA-DR/ CX3CR1 positivity in COVID-19 patients (FIG. 2). Indeed, CCR2 expression was reduced in the intermediate subset in COVID-19 patients.

**Figure 2.**
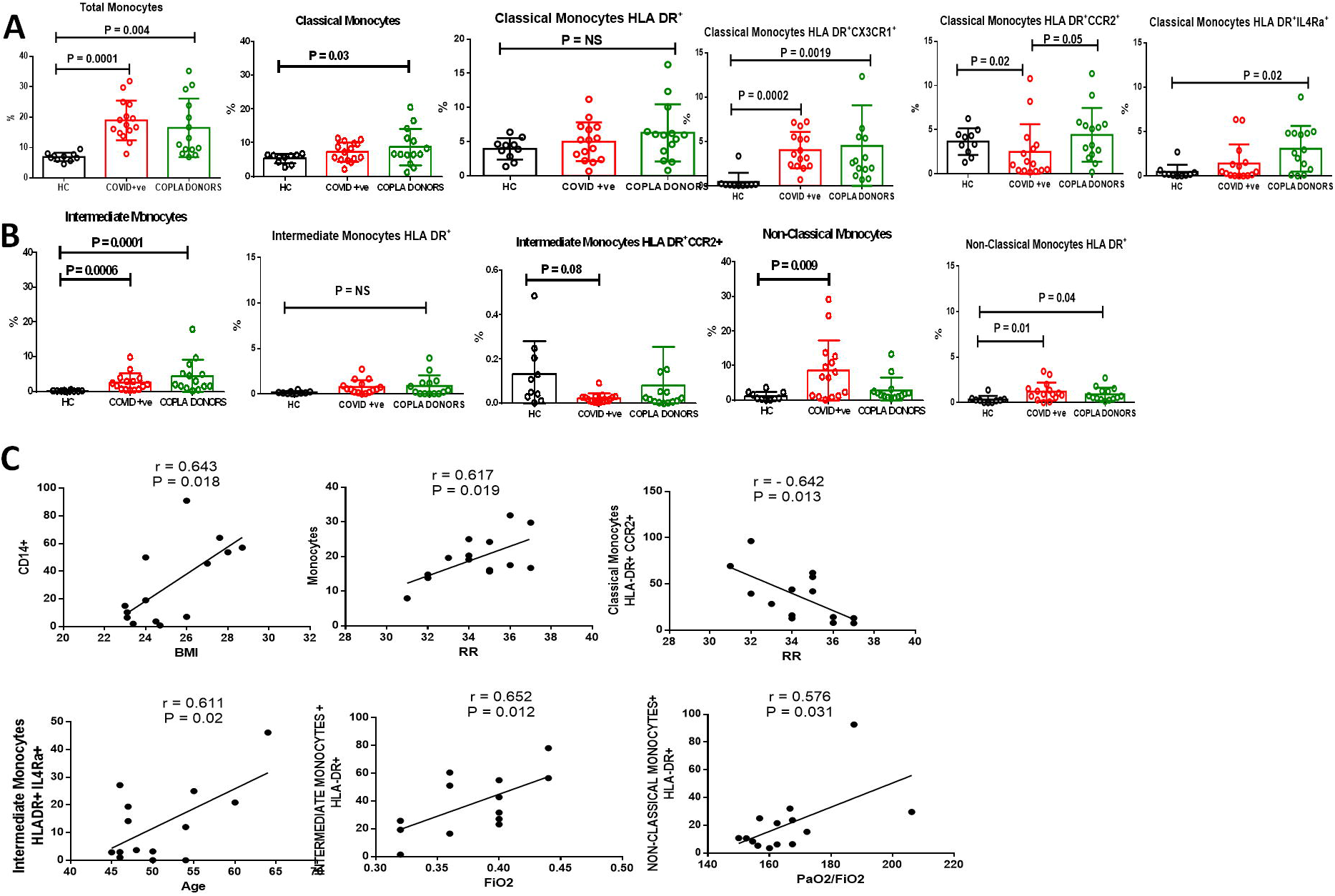
Total monocytes and their subset population in severe COVID-19 patients, recovered COPLA donors and healthy subjects. (**A)** Increased total monocytes in COVID-19 and COPLA donors, with enhanced expression of HLADR and CX3CR1 in classical monocytes. Copla donors also showed increased expression of CCR2 and IL-4Ra on HLADR+ classical monocytes. (**B)** Dot plots showing increased intermediate monocytes in COVID-19 and COPLA donors with respect to healthy subjects but with low expression of CCR2. COVID-19 patients showed increased non-classical monocytes and HLADR expression. (**C)** Increase in CD14+ expression and monocytes were significantly correlating with higher BMI and respiratory rate. Decreased CCR2 expression was also correlating with higher respiratory rate in COVID-19 patients. Increased HLADR on intermediate and non-classical monocytes and IL4RA expression was correlating with increased age, FiO2 and Pao2/FIO2levels. Error short bars in the graph shows the mean ± SEM. *P* < 0.05 is considered significant. r is representing correlation coefficient.

There was no significant difference in immune profile after a week in patients except classical monocytes increased in circulation (data not shown).

However, in COPLA donors, after three weeks of recovery, we have observed the classical monocyte compartment was significantly increased with no change in HLA-DR but with sustained expression of CX3CR1 (FIG. 2A-B).

In COVID-19 patients, increase in total monocytes were significantly correlated with increase in BMI and respiratory rates. However, intermediate monocyte was significantly increased with increasing age and Fio2 levels (FIG. 2C).

### Increased central memory T cells in COVID-19 Patients and COPLA donors

In patients, the CD3^+^ T cell compartment was not significantly different from healthy subjects, however, the ratio of CD4/CD8+T cells was skewed in patients towards more towards CD4^+^ T cells. T cells, when further were compartmentalized on the basis of CD45 RA, CCR7, and CD27expression, into naïve, effector, central memory and effector memory T cells, it was observed that in patients, CD4 naïve cells were reduced but central memory T cells was increased. However, in CD8 T cells compartment, naïve, effector, central, and effector memory subsets were increased compared to healthy (Fig. 3). COPLA donors did not show different numbers of CD4/CD8 T cells than healthy controls, however, they still had greater expression of effector and central memory but reduced effector memory compartment of CD8 T cells.

**Figure 3.**
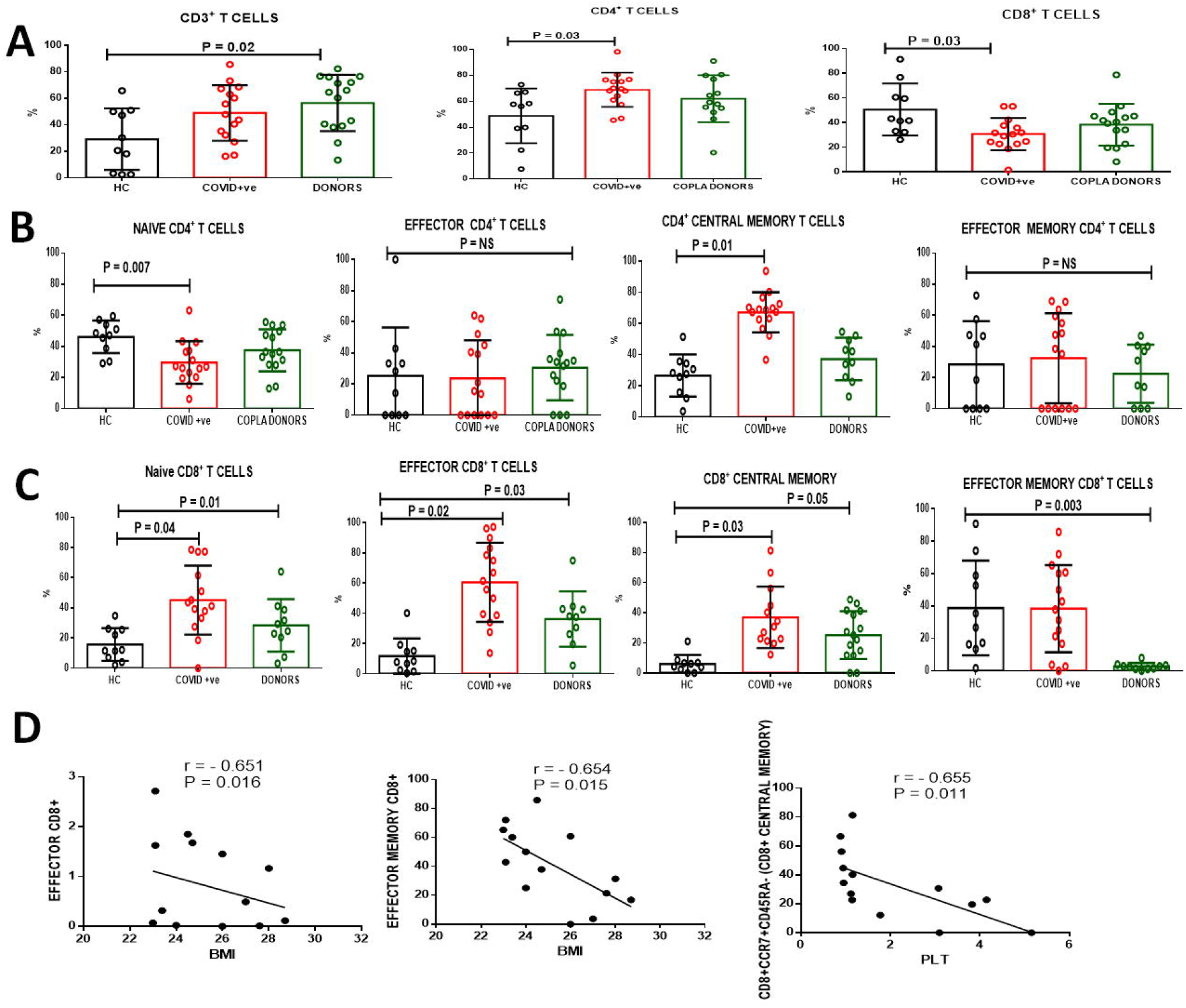
Total T cells and their subsets like naïve, effector and memory T cell populations in severe COVID-19 patients, recovered COPLA donors and healthy subjects. (**A)** Dot plot showing the percentage of circulating CD3, CD4 and CD8 T cell subsets. Error short bars in the graph shows the mean ± SEM. *P* < 0.05 is considered significant. **(B-C)** Percentage of CD4+ and CD8+T cell subsets as naïve, effector, central memory and effector memory three groups. **(D)** Decreased effector and effector memory CD8+ T cells was significantly correlated with higher BMI and decreased central memory cells were correlated with decrease in platelets in severe COVID-19 patients. *P* < 0.05 is considered significant. r is representing correlation coefficient.

In COVID-19 patients, increase in BMI and PLT were negatively correlated with effector, effector memory and central memory CD8^+^ T cells (Fig. 3D). We have also observed that in patients, increased effector CD8^+ve^ T but not CD4^+ve^Tcells could secrete IFN-γ, IL-17 and IL-2 upon LPS stimulation in vitro (Fig.4)

**Figure 4.**
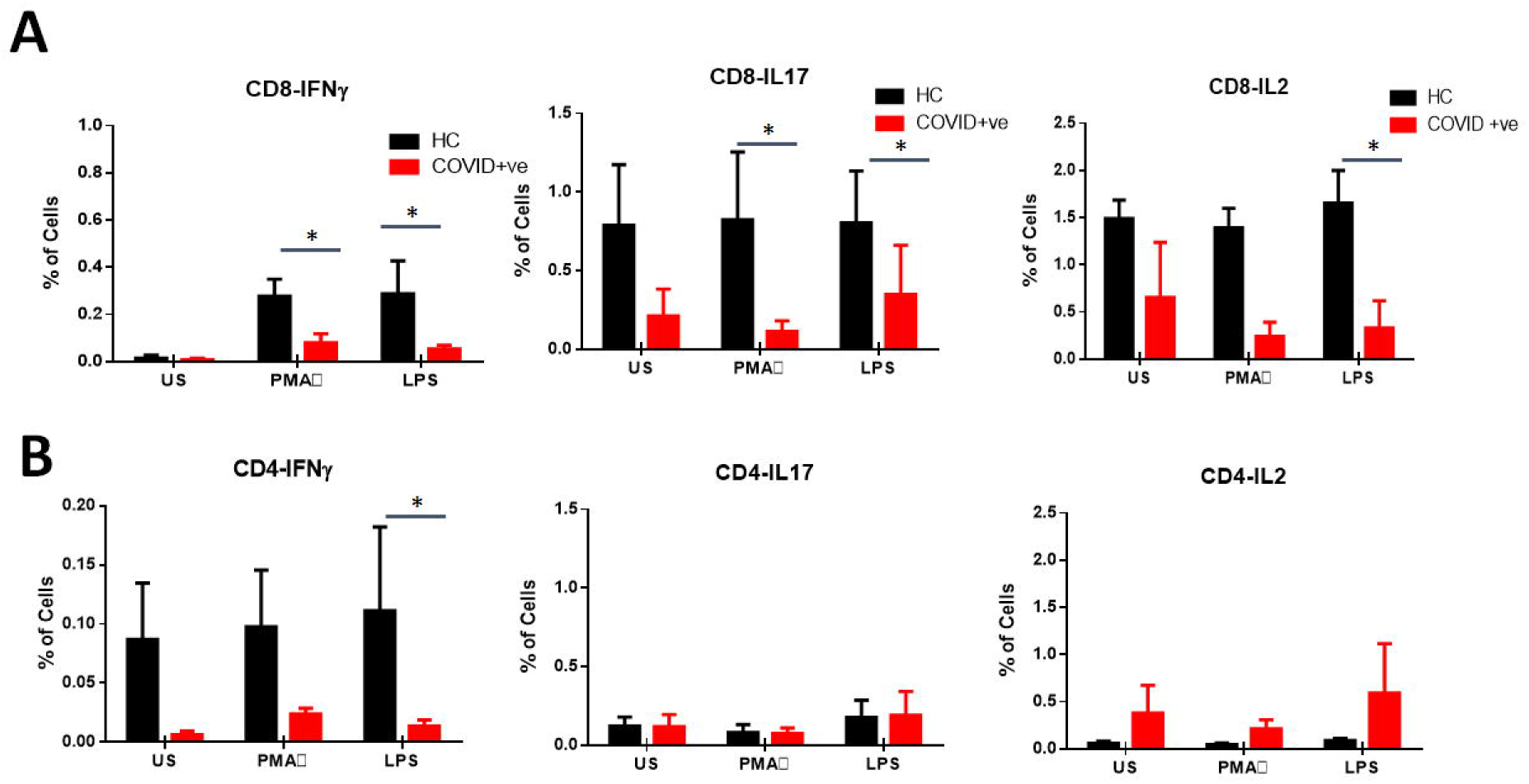
Functional assay of T cells displaying IFN-y, IL-17A and IL-2 secretory cytokines. PBMCs were stimulated with LPS and PMA / ionomycin as positive controls for 6 hours followed by phenotypic surface marker and intracellular staining. LPS Stimulated CD4/CD8 T cells were less poly functional in severe covid-19 patients. **(A-B)** Percentage of IFN-y, IL-17A and IL2 producing CD8 and CD4 positive T cells with or without stimulations. Abbreviation: US (Unstained), LPS (Lipopolysaccharides), PMA (phorbol 12-myristate 13-acetate). * *P* < 0.05 is considered significant.

### Downregulation of APRIL and BAFR in B cells

Total B cells were reduced in COVID patients, however, it was not significantly different from healthy subjects (Fig. 5). Gating strategies of B cell is given in supplementary material. But activated B cell number was decreased with lower expression of PDL1 and CD40. Indeed, it was observed that B cell-activating factor receptor (BAFFR) and a proliferation-inducing ligand (APRIL) expressing B cells were significantly reduced in both patients and COPLA donors (Fig.5 A-C). Correlation analysis also revealed decreased memory B cells, APRIL and BAFFR expression was strongly correlated with SOFA Score and age of COVID-19 patients (Fig.5 D).

**Figure 5.**
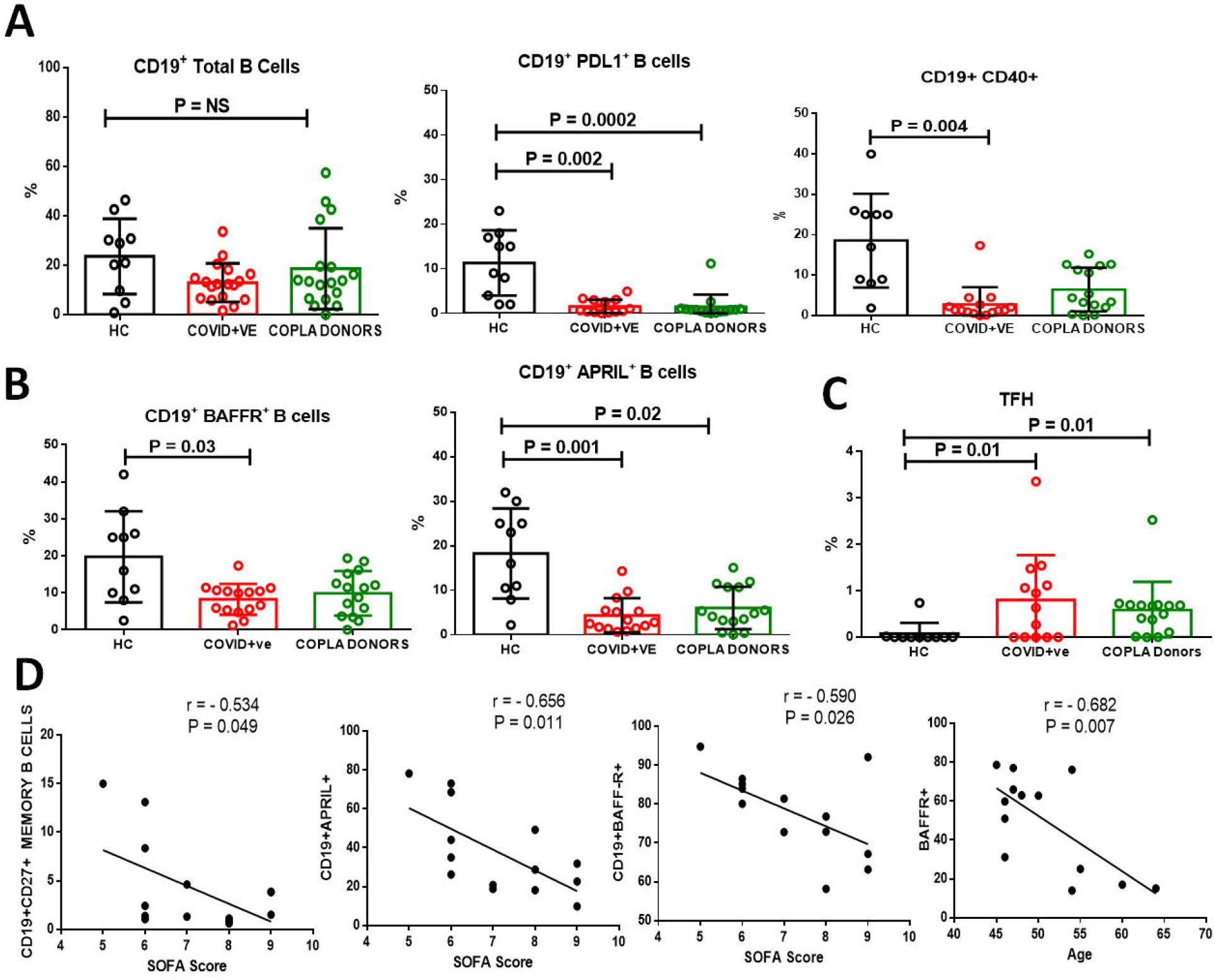
Cumulative dot plots showing the percentage of circulating B cells and its subsets and TFH cells in healthy subjects, severe covid10 and COPLA donors **(A)** Percentage of circulating CD19^+ve^ as B cells and expression of PDL1 and CD40on CD 19^+ve^ B cells. **(B)** Expression of BAFFR and APRIL on CD19 ^+ve^ B cells in all three groups. **(C)** Percentage frequencies of TFH cells in three groups.Error short bars in the graph shows the mean ± SEM. *P* < 0.05 is considered significant. **(D)** Decreased percentages of B cells correlating with increased SOFA score in severe COVID-19 patients. Decreased expression of APRIL and BAFFR on B cells correlating with increased SOFA score and age in severe COVID-19 patients. *P* < 0.05 is considered significant. r is representing correlation coefficient.

## Discussion

Analysis of plasma analytes using cytokine bead assay revealed the involvement of inflammatory monocytes by raising the levels of MIPs, MIF, and MCP1 in COVID-19 patients than healthy controls. Indeed, most of the pro-inflammatory cytokines such as IL-2, IL-1b, Il-6 IFN-γ, and IP-10 were expressed similar to the range in healthy controls. Only four patients who did not survive for a week had marginally increased IL-6 levels along with MIPs and MIF. This analysis suggests that other than IL-6 and TNF-α, other pro-inflammatory cytokine storms also do play a role in the severity of COVID-19. Earlier findings also showed high-levels of IP-10, MCP-1, MIP-1A, in most of the patients with severe COVID-19 (Prompetchara et al 2020). ONG et al 2020 have also revealed that instead of an increase in the typical pro-inflammatory cytokine storm, dynamic immune responses of Wuhan COVID-19 patients exhibited the role of IL-1 signalling in severe cases. We have also observed that IL-6 levels were significantly correlated with FIO2, It is also observed that, COVID-19 is more severe in elderly and subjects with comorbidities but also having scope of blunted immunity in elderly may reflect hypo responsive for cytokine storm.

Albeit, convalescent plasma donors even after three weeks of recovery had increased MMPs and MIPs but with a potential increase in MCSF and IL-18 (Fig. 1E) showing the existence of more inflammatory monocytes. Convalescent plasma donors had heightened immune responses even after three weeks of recovery but showed significant changes in IL-18, MCSF. Increased levels of IL-18 was very well considered beneficial for recovery of COVID-19 (Wen et al 2020).

This also suggests that patients who have recovered and considered for COPLA donations still have compromised immunity with sustained expression of inflammatory monocytes and activated T cells. The effects of these inflammatory cytokines should be observed post-infusion into plasma recipients suffering from COVID-19, and are already in a hyper inflammatory state.

SARS-COV2 may primarily infect monocyte and CD14+ subsets with higher expressions of CCR2 and CX3CR1 can differentiate and migrate from the periphery to the target sites as macrophages during infection (Ziegler-Heitbrock et al 2010). Monocytes with higher expression of CCR2 are also considered as senescent monocytes causing inflammatory disease in the elderly due to their shortened telomeres (Merino et al 2011). Although convalescent plasma therapy significantly reduced the respiratory rate, improved O2 saturation, and improved the PaO2/FiO2 ratio in COVID-19 patients. But donors had no significant decline in monocyte activation and differentiation.

Increased plasma monocyte chemotactic protein-1 (MCP-1) and CCR2 on monocytes in COVID-19 patients and COPLA donors strongly suggest that SARSCOV2 specifically target this population and is responsible for keeping sustained inflammatory environment even after clearance of this virus (Zhang et al 2020).

In viral infections, although the functional role of CD8+ T cell is crucial still needs to be well-modulated in order not to cause severe pathology. But T cell responses especially CD8+ T cells in SARS-CoV2 infection showed greater magnitude in the secretion of IFN-γ and TNF-α than CD4+ T cells. The early rise of CD8+ T cells and IL-4, IL-5, and IL-10 producing TH2 cells were detected more in patients with mortality. The present study revealed a bigger pool of CD8+T cells secreting IL-17, IFN-γ, and IL2 than CD4 T cells, however lower than healthy subjects. Although, the protective or destructive role of Th17 in human coronavirus infection remains unanswered (ONG et al 2020). Indeed in the convalescent phase, we observed Th1 type helper T cells, but still, there was a low expression of activation markers on B cells. A proliferation induced ligand (APRIL) and B cell-activating factor receptor (BAFFR) expression regulate the compartment of translational B cells to mature B cells. Recently, APRIL and BAFF expression was also observed more in IL-10 producing regulatory B cell compartment. Bregs acts as co-partners with Tregs in inhibiting excessive inflammation. Through, Bregs skew T cells expression and also functional in suppressing Th1 and Th17 differentiation, but in our study we have observed low expression of BAFFR and APRIL on B cells suggesting that B cells were not actively proliferating, although in COPLA donors their expression was enhanced better than COVID patients, still it was under-expressed than healthy.

In conclusion, SARS CoV-2 infection induces pathogenesis by monocyte trafficking and differentiation via CCR2 and CX3CR1, MIPs, and MIFs and with reducing APRIL and BAFFR expression, and there was sustained expression of these markers even after three weeks of recovery in COPLA donors.

## Materials and Methods

### Patients and Subjects

We prospectively recruited SARS-CoV-2 positive (by real-time PCR assay) severe COVID-19 patients (n=34) at the Department of Internal Medicine, Lok Nayak Hospital, a designated COVID-19 treatment centre in New Delhi and COVID-19 recovered patients who have recovered from mild disease and considered for convalescent plasma donation after three weeks of their recovery (n=15), at Institute of Liver and Biliary Sciences (ILBS), from May-July 2020. Severity of disease was characterised by respiratory distress, respiratory rate (RR) ≥30/min, oxygen saturation level less than 93% in resting state, the partial pressure of oxygen (PaO2)/oxygen concentration (FiO2) ≤ 300 mmHg, lung infiltrates > 50% within 24 to 48 hours] (Table 1). Patients less than 18 years or more than 65 years of age, those with co-morbid conditions(cardiopulmonary disease, structural or valvular heart disease, coronary artery disease, COPD, chronic liver disease, chronic kidney disease), patients presenting with multi-organ failure, pregnant females, individuals with HIV, viral hepatitis, cancer, morbid obesity with a BMI>35 kg/m2, extremely moribund patients with an expected life expectancy of <24 hours, or failure to obtain informed consent were excluded from this study.

Recovered patients were symptom free for at three weeks and had negative real-time PCR assay. To compare the intensity of immune markers healthy subjects (n=10) were enrolled in this study. The study was performed in accordance with Institutional Ethical Committee approval (No.F.37/(1)/9/ILBS/DOA/2020/20217/260).

### Sample Collection

Both nasopharyngeal and oropharyngeal swabs were taken in a 3 ml viral transport media. 6-8 ml peripheral blood was collected in EDTA coated vacutainers from COVID-19 patients at the time of admission and recovered patients after 14 days of their recovery at the time of convalescent plasma donations. Plasma was separated and stored at -80°C for further use.

### Performance of RT-PCR

A volume of 200 µl of the sample was further processed for viral nucleic acid extraction by Qiasymphony DSP Virus/ Pathogen mini kit (Qiagen GmbH, Germany) as per the manufacturer’s protocol.

The extracted elutes of RNA was subjected to RT-PCR for the qualitative detection of both E as well as ORF 1ab (RdRP) genes of SARS-CoV-2 virus using a commercial RT PCR kit (nCoV RT–PCR, SD Biosensor, Gyeonggi-do, Republic of Korea) as per the kit literature. The sample was considered positive when fluorescence was seen in both the target genes E as well as RdRP up to a cycle threshold (Ct) value 36. Ct value for each gene was noted separately and keeping in mind that there exists an inverse relationship between Ct value and the amount of viral nucleic acid in the specimen, Ct value was utilized as a marker to monitor the clinical progress of the patients.

Two consecutive negative test results of Real-time reverse transcriptase Polymerase chain reaction (RT-PCR) were done 24 hours apart from combined oral and nasopharyngeal swab for SARS CoV-2 for donation consideration.

### Demographic Data and Laboratory Parameters

We collected demographic data, symptoms, and clinical signs from the medical records. Laboratory tests were conducted for serum protein, CBC; transfusion-transmitted infections (hepatitis B virus, hepatitis C virus, HIV, malaria, and syphilis), blood grouping, and IgG antibody against SARS CoV-2.

### Analysis of Plasma analytes using Bead Assay

The concentrations of twenty one plasma cytokines, chemokine and growth factors such as IL-1β, IL-2, IL-18, IL-10, IFN-γ, IL-6, TGFb1, MIF, MIP-1α, MIP-1β, MIP-3α, ITAC, MCP1, FRACTALKINE, ENA78, IP-10, MCSF, LEPTIN, VEGF-A, MMP12 and E SELECTIN were determined in 25-30 μl of plasma in all three groups by using multiplexed procataplex cytokine bead assay (Thermo Fisher Scientific, Bender MedSystems GmbH, Campus Vienna Biocenter 2, Vienna Austria) following the complete details on Luminex xponent 3.1TM Rev. 2 (Luminex Corporation, 12212 Technology Boulevard, Austin, Texas, USA). Standard curve was drawn using standards provided in the kit. Values in samples were determined corresponding to the standard curve drawn. The lower limits of detection of the test for each cytokine is given in the supplementary data.

### Multiparametric Whole blood Immunophenotyping

Monocytes, T-cells, B cells, and Tregs were characterized in whole blood. 100 µl of whole blood sample was stored in 700 µl of FACS Lysing solution (BD Pharmingen, USA) and was stored in -80□C for further use. Using specific antibodies against surface and intracellular markers labelled with different fluorochromesblood mononuclear cells were characterized.

All samples were processed on single day in one batch to avoid any batch to batch processing variation. FACS Lysed samples were thawed in 37□C water bath for a minute and immediately centrifuged and washed twice with 1x PBS containing 0.5% BSA with 0.1% Sodium Azide at 400g for 3 minutes at room temperature. After two washes, cell pellet was stained for surface expressing markers by incubating with appropriate cell specific antibodies for 30 minutes at room temperature in the dark. After incubation, cells were washed with 1x PBS containing 0.5% BSA with 0.1% Sodium Azide and spun at 400g for 3 minutes to discard the supernatant. Collected pellet was permeabilized with 200 µLFACS permeabilisation solution (BD Pharmingen, USA) and incubated for 10 minutes before washing the cells with 1x PBS containing 0.5% BSA with 0.1% Sodium Azide. Washed cells were centrifuged and collected cell pellet was further incubated for 30 minutes with specific antibodies for intracellular expression of markers and washed with 1xPBS. Antibodies were labelled with different flouorochromes and used **for B Cells**; anti-CD3, CD14, CD56, CD19, CD27, CD21, IgD, CD40, CD38, CD274, CD185, CD268, BAFFR and APRIL, **For monocytes;** anti-CD3, CD56, CD19, CD14, CD16, HLA-DR, CD11b, CD11c, CCR2, CX3CR1, CD80, CD124, CD45, CD163, CD71, CD206,CD123, CD68, and **for T cells and its subsets**; anti-CD3, CD4, CD25, CD27, CCR7, FOXP3, GATA3, CD194, CD278-, CD45RA, CD127. Finally cell pellet was acquired on FACS ARIA analyzer cum sorter and a minimum of 100,000 events were analyzed by using FlowJo software version10.2.

### T cells Functionality

To investigate the CD4 and CD8 T cell functionality in COVID-19 positive patients, PBMCs were cultured in 96 well plate in RPMI media containing 10% FBS, with or without PMA / Ionomycin (positive control) (PMA 2 ng/mL; ionomycin 1 μg/mL; Merck, Darmstadt, Germany) and 10 µg /ml LPS (Merck, Darmstadt, Germany) at 37°C, 5% CO2 for 6-7 hours. After 1 hour of incubation, 1 μg/mL brefeldin A (BD Pharmingen, USA) was added to all the wells. After incubation, cells were surface stained for 25-30 minutes with anti-CD3FITC, anti-CD8BV421 and anti-CD4 PE/cyanine5.5 (Cy5) followed by 10 minutes permeabilization with 100 µL of permeabilising solution(BD Biosciences, USA) and washed twice with 500 µL 1x cytoperm. After washing, cells were stained for intracellular expression of IL-2, IFN-γ and IL-17 with anti-IL2 APC, anti-IFN-γ PE, anti-IL-17 PE-Cy7 (BD Biosciences and Pharmingen, USA), incubated for 25-30 minutes followed by washing with PBS and fixing the cells in 0.1 % PFA before acquiring on a BD Verse flow cytometer. Data was analyzed using FlowJo software version 10.

### Statistical analysis

Continuous variables were expressed as mean (SD) or median (range) and compared by student T-test or Mann-Whitney U test as appropriate. The categorical data were analyzed using Chi-Square or Fisher’s exact test. To compare pre- and post-values, a paired t-test or Wilcoxon signed-rank test was used. Pearson correlation was calculated with respect to many clinical parameters.

## Data Availability

Yes, I agree

## Abbreviations

COPLA donors: (Convalescent Plasma donors)
HC: (Healthy Controls)
APRIL: (A proliferation-inducing ligand)
BAFFR: (B-cell activating factor receptor)
(DCs): Dendritic cells
PCR: (Polymerase Chain Reaction)
IL: (Interleukins)
IFN: (Interferon)
TGF-b: (Transforming growth factor b)
MIP: (macrophage inflammatory protein
MCP: (Monocyte chemoattractant protein
ITAC: (interferon-inducible T-cell α chemoattractant)
ENA78: (neutrophil-activating peptide)
VEGF: (vascular endothelial growth factor)
IP-10: (IFN-γ inducible protein 10)
MMP: (Matrix metalloproteinase)
RR: (respiratory rate)
PaO2: (Partial pressure of oxygen)
FiO2: (fraction of inspired oxygen)
COPD: (Chronic obstructive pulmonary disease)
BMI: (Body Mass Index)
CBC: (Complete blood count)
HIV: (Human Immunodeficiency virus)
PBMC: (peripheral blood mononuclear cells)
(WB): Whole blood
(SD): Standard deviation
SOFA score: (Sequential Organ Failure assessment score)

## Acknowledgments

We thank our technician Mr. Dileep at MCM for the storage and initial processing of samples. The study was supported by intramural funding of the corresponding author.

## Author contributions

MB, SK SKS, and AK recruited, characterized, and conducted the clinical investigations of all patients and COPLA donors. RS and NTP processed all samples in the laboratory. NTP conceptualized the study and wrote the manuscript. SKS provided critical inputs. JK and IM conducted the cytokine bead assay. SA, PY, GR, JS, and NTP analysed all the data of cytokine bead assay and flow cytometry.

