## Supplementary information for "Sustained expression of inflammatory monocytes and activated T cells in COVID-19 patients and recovered convalescent plasma donors"

| **Cytokine** | **Pg/ml** |
| --- | --- |
| IL-6 | 10 |
| IL-2 | 5.71 |
| IL-10 | 1.7 |
| IFN- γ | 15 |
| MIF | 0.21 |
| MCP-1 | 3.93 |
| IP-10 | 2.03 |
| IL-1b | 1.99 |
| IL-18 | 17 |
| TGF-b1 | 3 |
| MIP-1α | 2.04 |
| MIP-1β | 5.64 |
| MIP-3α | 4.44 |
| ITAC | 3.74 |
| FRACTALKINE | 1.25 |
| ENA78 | 6.59 |
| MCSF | 14 |
| LEPTIN | 16 |
| VEGF-A | 5.37 |
| MMP12 | 5.76 |
| E SELECTIN | 206 |

**Supplementary Table: Lower limit of Cytokine levels in Cytokine Bead Assay**

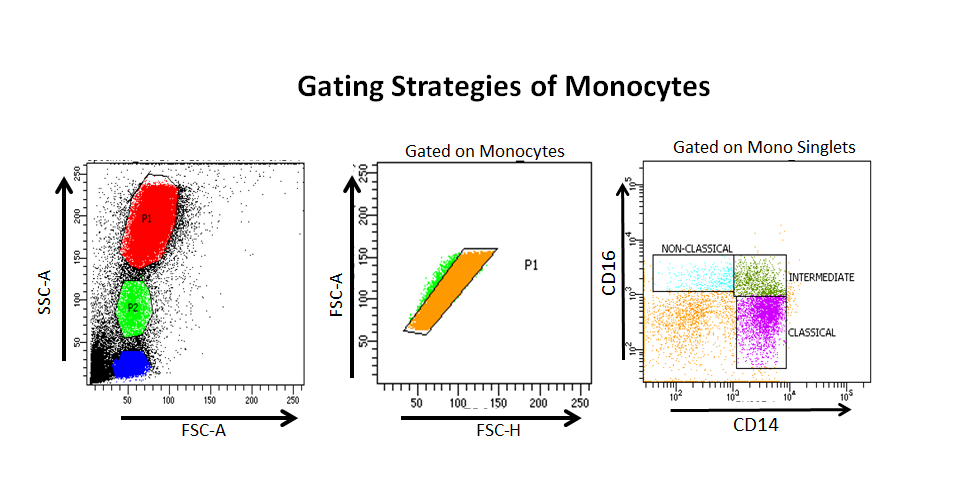

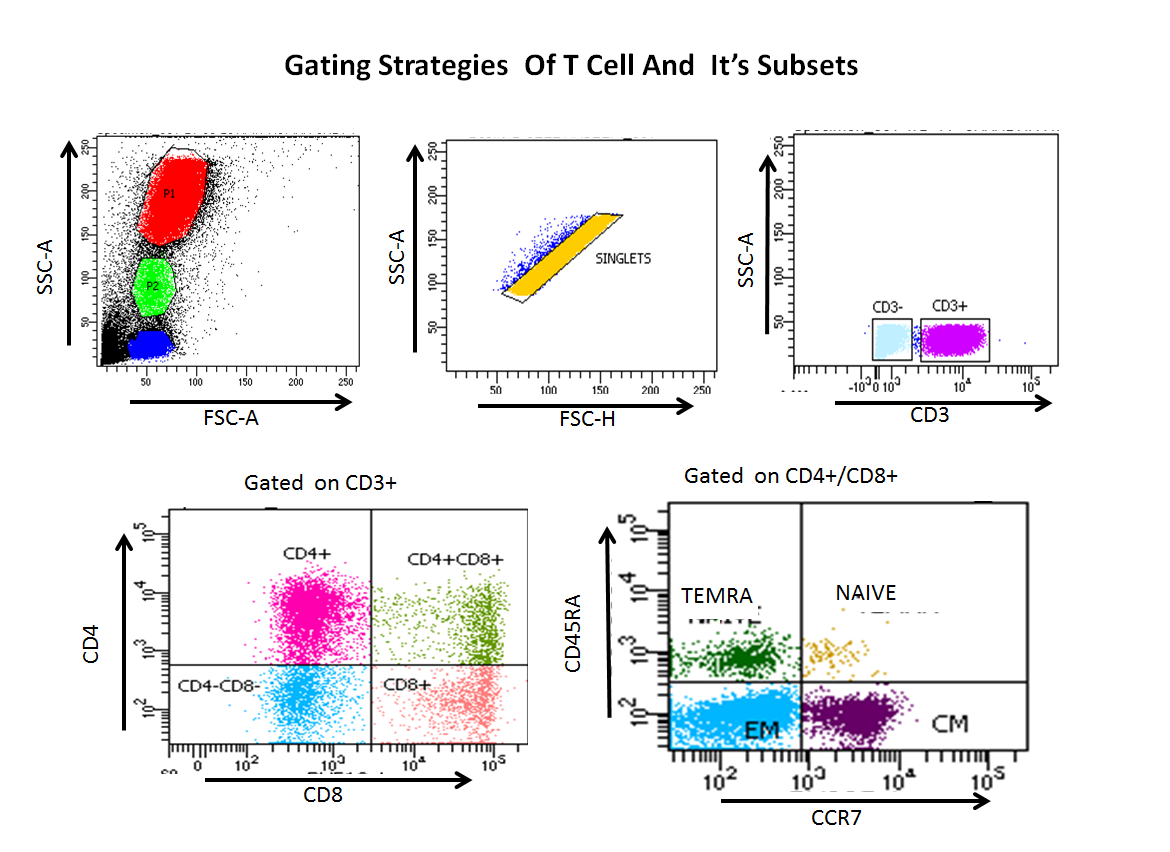

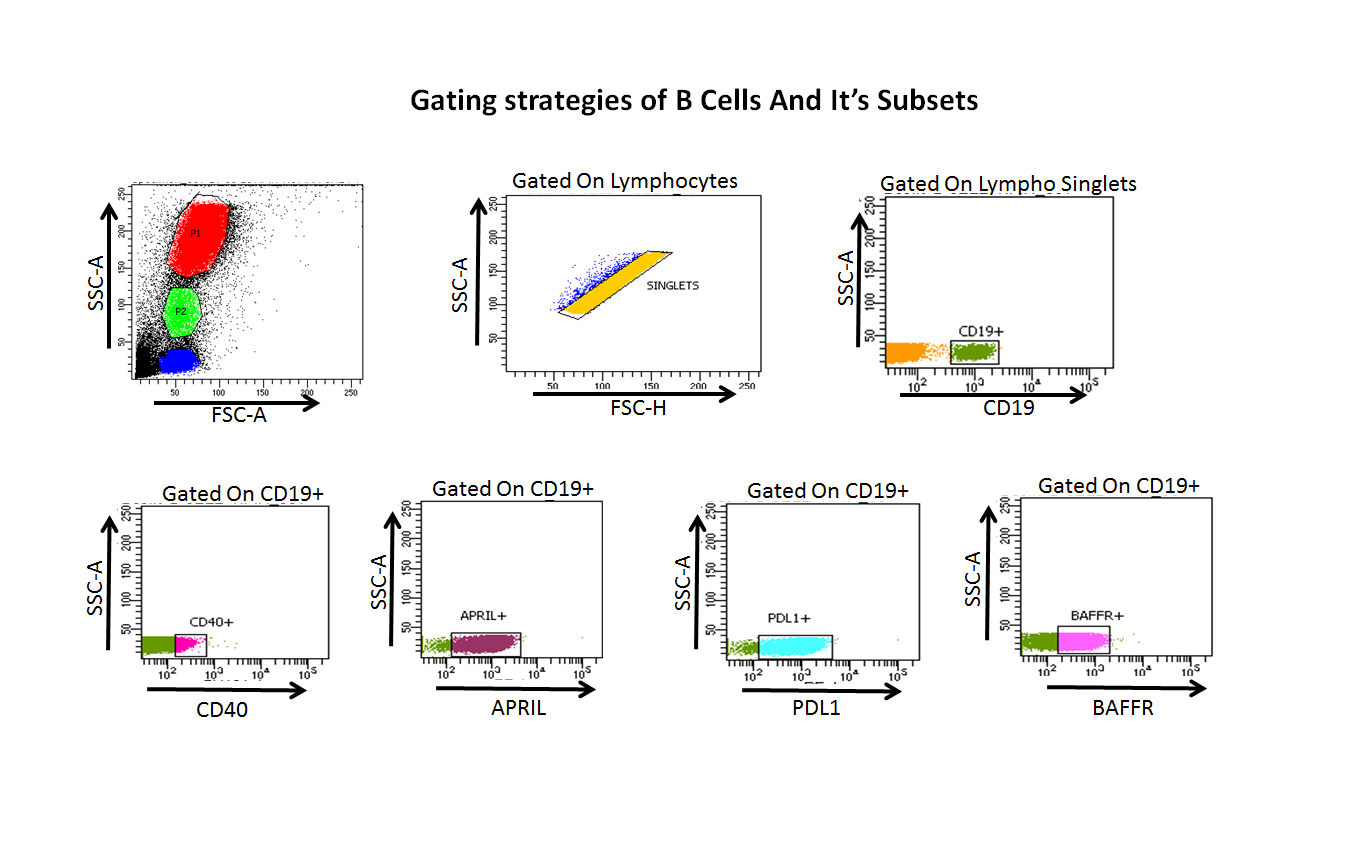
